# Automatic multi-object organ detection and segmentation in abdominal CT data

**DOI:** 10.1101/2020.03.17.20036053

**Authors:** Oliver Mietzner, Andre Mastmeyer

## Abstract

The ability to generate 3D patient models in a fast and reliable way, is of great importance, e.g. for the simulation of liver punctures in virtual reality simulations. The aim is to automatically detect and segment abdominal structures in CT scans. In particular in the selected organ group, the pancreas poses a challenge. We use a combination of random regression forests and 2D U-Nets to detect bounding boxes and generate segmentation masks for five abdominal organs (liver, kidneys, spleen, pancreas). Training and testing is carried out on 50 CT scans from various public sources. The results show Dice coefficients of up to 0.71. The proposed method can theoretically be used for any anatomical structure, as long as sufficient training data is available.

## 1 Introduction

Virtual reality (VR) based simulation of needle interventions for training and planning is slowly gaining importance in clinical teaching and routine. VR methods can be used for various tasks, ranging from training scenarios for the medical student and staff, to individual patient related simulations [7, 11, 15, 8, 9, 3, 16] of planned operations [6, 12, 14, 13, 10]. The necessary individual patient models should be available fast and be accurate to guarantee a plausible simulation.

The first step in producing patient models is to acquire patient image data. This data will be used for the following steps in the modeling process. It is desirable to use high-quality image data, because inaccuracies will be carried over to the resulting model. The next step is the coarse localisation of the organs inside organ-specific volumes of interest (VOI) to simplify their subsequent segmentation.

The aim of our study, is to automatically detect bounding boxes for organs in abdominal CT data by a learning-from-example method. The detected bounding boxes are then used for segmentation. Detection of the abdominal organs can be a challenge, because of the variety of shapes these organs can have. The intensity based features of these organs are also challenging, because the intensity range of neighbouring organs often overlaps. We detected five abdominal organs (liver, right kidney, left kidney, pancreas, spleen), that are commonly used in simulations.

We used a random-forest method, that is developed by Criminisi et al. [1] to automatically detect the bounding boxes of abdominal organs. The method uses random-regression-forests to predict the location of organ bounding boxes in CT data. Furthermore, we used the 2D U-Net, proposed by Ronneberger et al. [18], to automatically segment organs. The training and testing is carried out on a database of 50 abdominal CT scans, that had to be segmented beforehand.

## 2 Material and Methods

The database, that is used for training and testing, consisted of 50 CT scans from different sources. The data sets contained only abdominal CT data. The sets also varied in quality and field of view, to capture the variability of organs and scans. The scans are not only different in the number of slices (64-861), thickness of slices (1-5 mm) and field-of-view, but also in image noise. This variability in the datasets is important, to ensure robustness vs. typical inference factors of clinical image data during training and application. All five target organs (liver, pancreas, left kidney, right kidney, spleen) are included in the scans.

Only a small fraction of the segmentation maps contained all five target organs, thus manual structure segmentation was necessary frequently.

### 2.1 Definition of ground truth bounding boxes

The segmentation maps associated to the CT scans are the basis for the extraction of ground truth bounding boxes to be learned. A bounding box can be described as a cubical polyhedron that completely encloses an object.

The corresponding organ bounding boxes of each of the scans can be created by scanning the segmentation maps for coordinate direction extremes of label occurrence. To create a three-dimensional bounding box, for each coordinate direction (*x, y, z*), we iterate slice-wise through the segmentation map and save the limits from an object label.

### 2.2 Training of the models

Our method is composed of two different machine learning approaches, random regression forests (RRF) and 2D U-Nets. The RRF’s are used to detect organ bounding boxes in CT data. The 2D U-Nets make use of the detected bounding boxes and learn the axial slices of each organ contained in them.

#### 2.2.1 Training random regression forests for bounding box detection

The decision trees used in RRFs split by minimising variance, then each leaf node outputs the mean of all label values in the node. We use RRFs to determine the location and extent of abdominal organs [1]. The main difference between random classification forests and RRFs, is the type of output that is predicted. While classification forests try to categorize objects, regression forests predict continuous values. Regression forests partition the data into manageable chunks to predict average values. As seen in Fig. 1, the method expects CT scans and ground truth bounding boxes as input.

**Figure 1:**
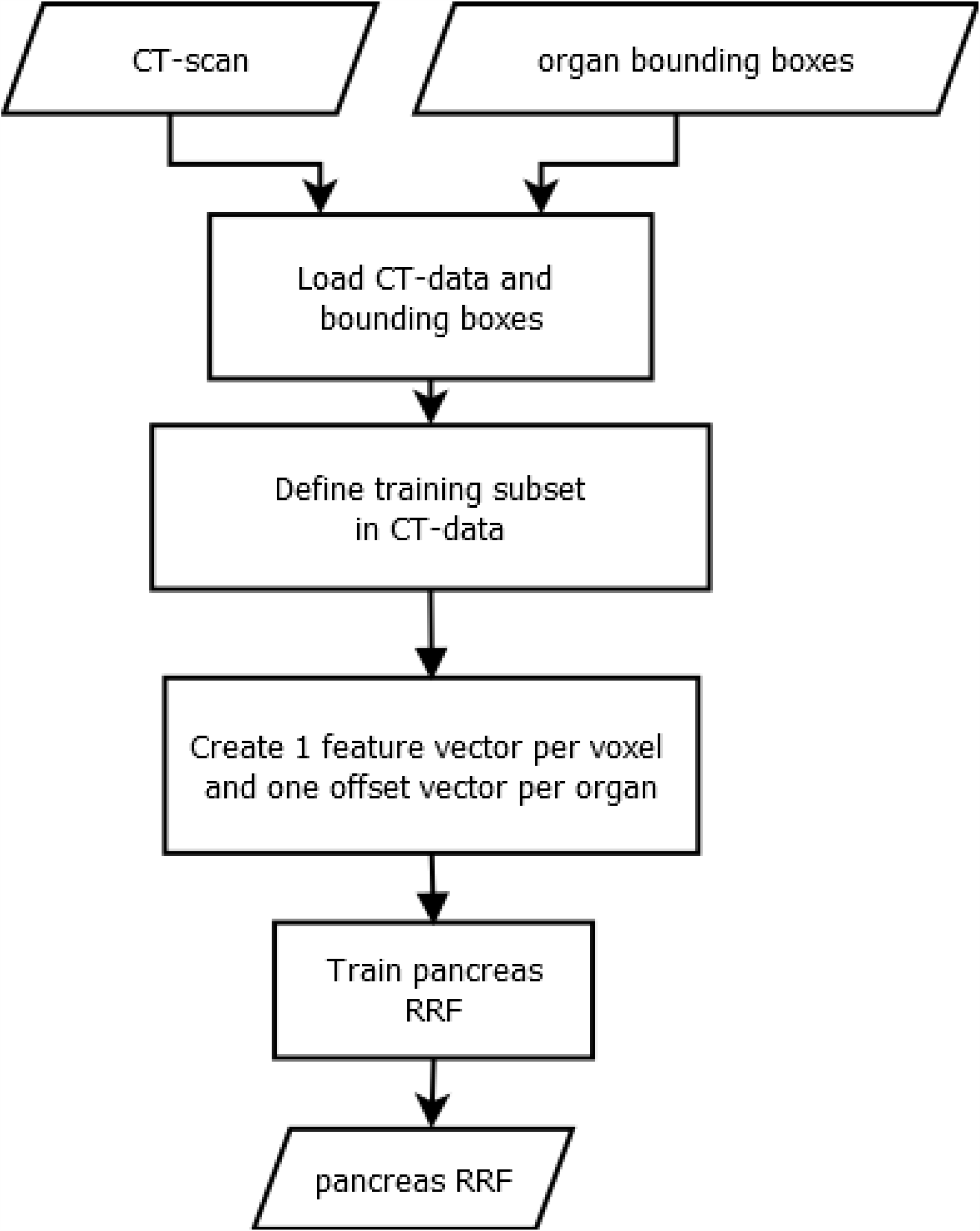
Training of a RRF: The inputs for the training process are CT scans and ground truth bounding boxes of the targeted organs. We create one feature vector and one offset vector for each voxel that is part of a predefined medial cylinder subset in the scan [1]. The trained RRF is able to predict the offset between a voxel and an organ bounding box.

A three-dimensional bounding box *b*_*c*_ of an organ *c* can be described by using a six-dimensional vector:

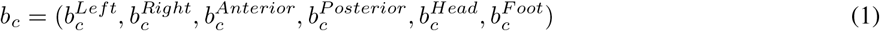

with coordinates in mm [1]. We run over all voxels *p* = (*x*_*p*_, *y*_*p*_, *z*_*p*_), which are within a specified radial distance (r = 5 cm) from the scan medial axis. The distance *d* between such a voxel and each of the bounding box walls, can be calculated by using *d*(*p*) = (*x*_*p*_ *− x*^*Left*^, *x*_*p*_ *− x*^*Right*^, *y*_*p*_ *− y*^*Anterior*^, *y*_*p*_ *− y*^*Posterior*^, *z*_*p*_ *− z*^*Head*^, *z*_*p*_ *− z*^*Foot*^) and is saved as the offset-vector to be learned. In contrast to Criminisi et al. [1], we use only 50 feature boxes, that are evenly distributed on three spheres (r = 5 cm, 2.5 cm, 1.25 cm) to generate the input feature vector. The feature boxes *F*_*j*_ are intended to capture the spatial and intensity context of the current voxel. For this purpose, the mean intensities of the feature boxes are calculated and saved in the feature vector. An example feature box is shown in Fig. 2. While training, the RRF learns the distance vector (later: output) to the reference bounding box using the feature boxes.

**Figure 2:**
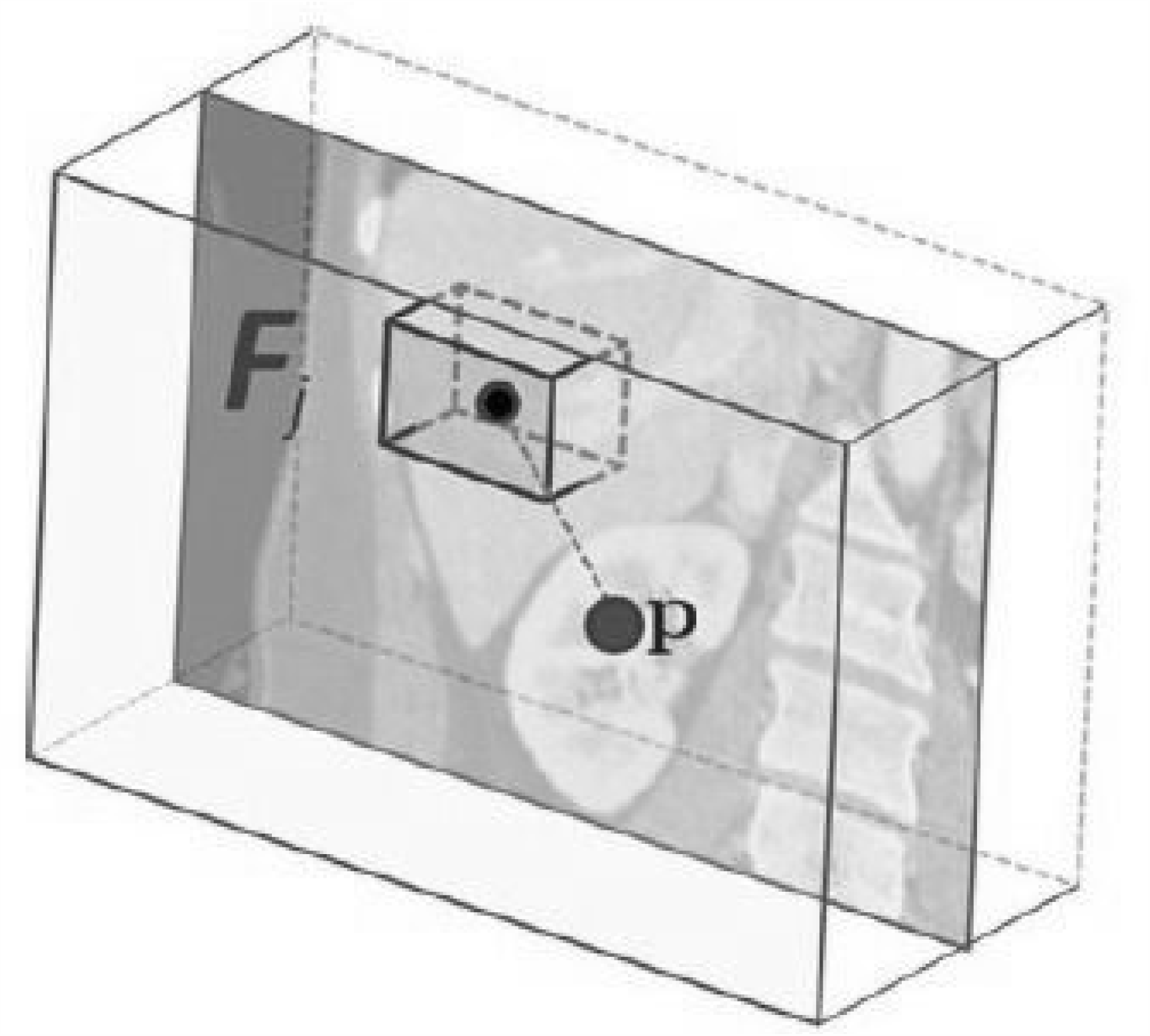
Example feature box: The feature box *F*_*j*_ is generated in correlation to the current voxel and calculated the mean value of a 3D image section [2].

#### 2.2.2 Training of a 2D U-Net for semantic segmentation

The training data for our U-Net consists of the expert segmentations and ground truth bounding boxes, as schematised in Fig. 3. The bounding boxes (VOIs) are then used to locally extract the intensity and label data from the CT scans and their corresponding label maps. As input, the 2D U-Net receives a VOI from the intensity data, while the same region within the label data is connected to the output. The VOIs contents are trained slice-wise axially. We use the 2D U-Net architecture proposed by Ronneberger et al. [18], which consists of nine layers with four down- and up-scale steps. The network was trained using batches of size 15 over 50 epochs. In addition, Adam optimization and a cross entropy loss function were used. We trained one U-Net for each organ, using a ReLU activation function.

**Figure 3:**
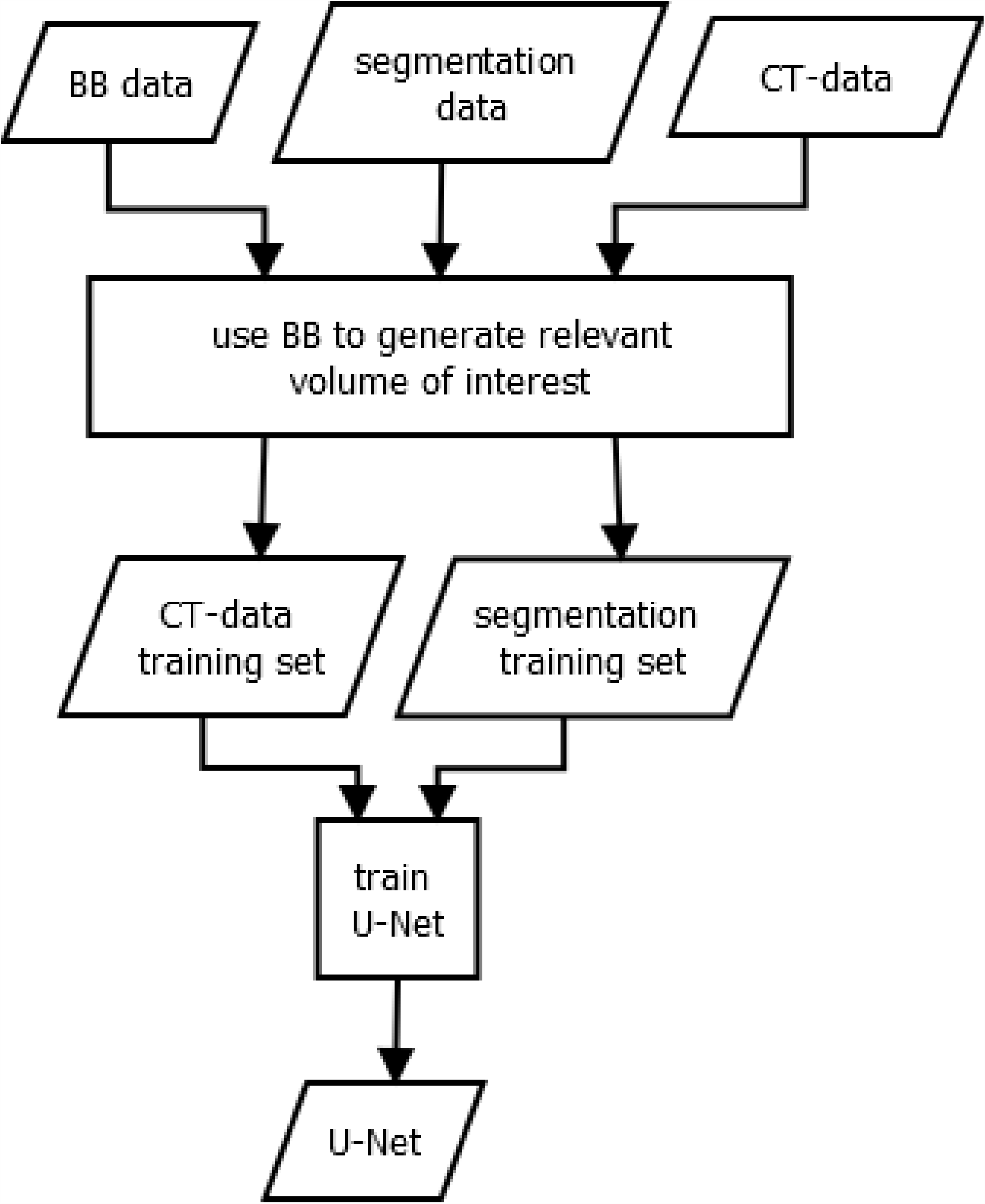
Training of a U-Net: The inputs for the training process are ground truth bounding boxes (BB), CT scans and their corresponding segmentation maps. The box is applied to the CT- and segmentation data to extract the relevant image region. Inside the organ VOIs, the segmentation is learned. The process results in an organ-wise U-Net, that can segment image regions.

### 2.3 Application of the models

In the first step, the organ specific RRFs predict the organ bounding box candidates. Then, a distance vector is selected by majority voting and converted into a six-dimensional vector to describe the final organ-specific bounding box. Now, the 2D U-Net model gets the corresponding bounding box as input. Since the 2D U-Net only accepts a static number of voxels, the BB is resized to a fixed size. The 2D U-Net uses the data contained inside the given bounding box, to segment the corresponding organ iteratively for each axial slice. The output is a 3D segmentation map of the full target organ.

### 2.4 Evaluation

We used a five-fold Monte Carlo cross-validation based on a 30:20 (train:test) data split. The target bounding boxes extend along the three axis *x, y, z*. Based on the proposition by [1], the quality, of the prediction can be determined, by measuring the distance, between the position of the detected bounding boxes and the position of the ground truth bounding boxes. The distance is equal to the absolute value of the difference *E* between a ground truth coordinate *GT* and a predicted coordinate *Pred*, given in (2).

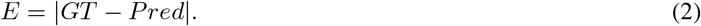

The resulting segmentations of the combined application are compared against the reference segmentations by using the Dice coefficient [19].

## 3 Results and Discussion

Table 1 shows the bounding box localisation error in ṁm, for our version of the method proposed in [1]. We compared our results to the results in [1], except the pancreas, which was not examined by Criminisi et al. [1].

**Table 1:**
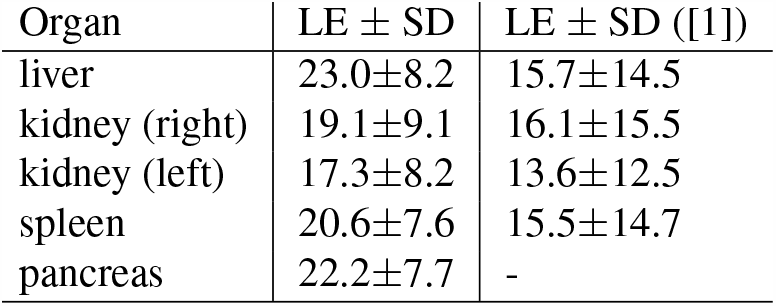
Comparison of bounding box localisation error (LE) in mm, for our version of the method proposed in [1].

| Organ | LE ± SD | LE ± SD ([1]) |
| --- | --- | --- |
| liver | 23.0±8.2 | 15.7±14.5 |
| kidney (right) | 19.1±9.1 | 16.1±15.5 |
| kidney (left) | 17.3±8.2 | 13.6±12.5 |
| spleen | 20.6±7.6 | 15.5±14.7 |
| pancreas | 22.2±7.7 | - |

All five organs produced similar results. The left kidney achieves the best value with only 17.3 mm difference between predicted and ground truth bounding box, while the liver produced the highest results with 23 mm. The overall detection accuracy could not reach the results of [1], with results that deviated with up to 6.3 mm. Nevertheless the standard deviation (SD) was lower than in [1]. The results could also be calculated for each individual axis. This showed that the *z*-axis produces the biggest errors.

The boxes we estimated are comparable to the results shown in [1]. The loss in accuracy can be accounted to a fewer number of training samples. The use of a wider variety of scans may help to detect organs with varying forms. While the extent of the kidneys and spleen were similar in different patients, the extent of the liver and pancreas may vary, resulting in a bigger localisation error. Especially the prediction along the *z*-axis is not accurate enough and resulted in boxes that are displaced. This can be attributed to deviations in the resolution of the scans. Due to misalignment, some segmentation masks are cut off, leading to parts of some organs missing. The standard deviations are too high, indicating highly varying results.

Table 2 shows the Dice coefficients achieved by our method for all five target organs. We compared our values to studies, that tried to segment the same organs with automatic methods.

**Table 2:** Comparison of [3] mean - over all per organ results - Dice coefficients and standard deviation (SD) of our general method vs. other one organ-focused methods.

| Organ | Dice ± SD | Other Dice Coefficients |
| --- | --- | --- |
| liver | 0.71±0.31 | 0.97 [17] |
| kidney (right) | 0.55±0.34 | 0.91 [5] |
| kidney (left) | 0.67±0.32 | 0.91 [5] |
| spleen | 0.71±0.26 | 0.94 [4] |
| pancreas | 0.32±0.28 | 0.74 [4] |

The best results show up for the liver and spleen. Both organs achieve a dice coefficient of 0.71. The segmentation of the pancreas achieved the lowest dice coefficient with only 0.32. The standard deviation is similar for all organs ranging from 0.26 to 0.34. Varying results can be attributed to different target organs. Especially the pancreas is a challenge, because of its shape and intensity similarity to surrounding tissue. The right kidney is slightly worse vs. left. This could be attributed to the also larger localisation error from Table 1. The position errors of the right kidney BBs seem to be affected by the vicinity of the liver. An example segmentation is presented in Fig. 4.

**Figure 4:**
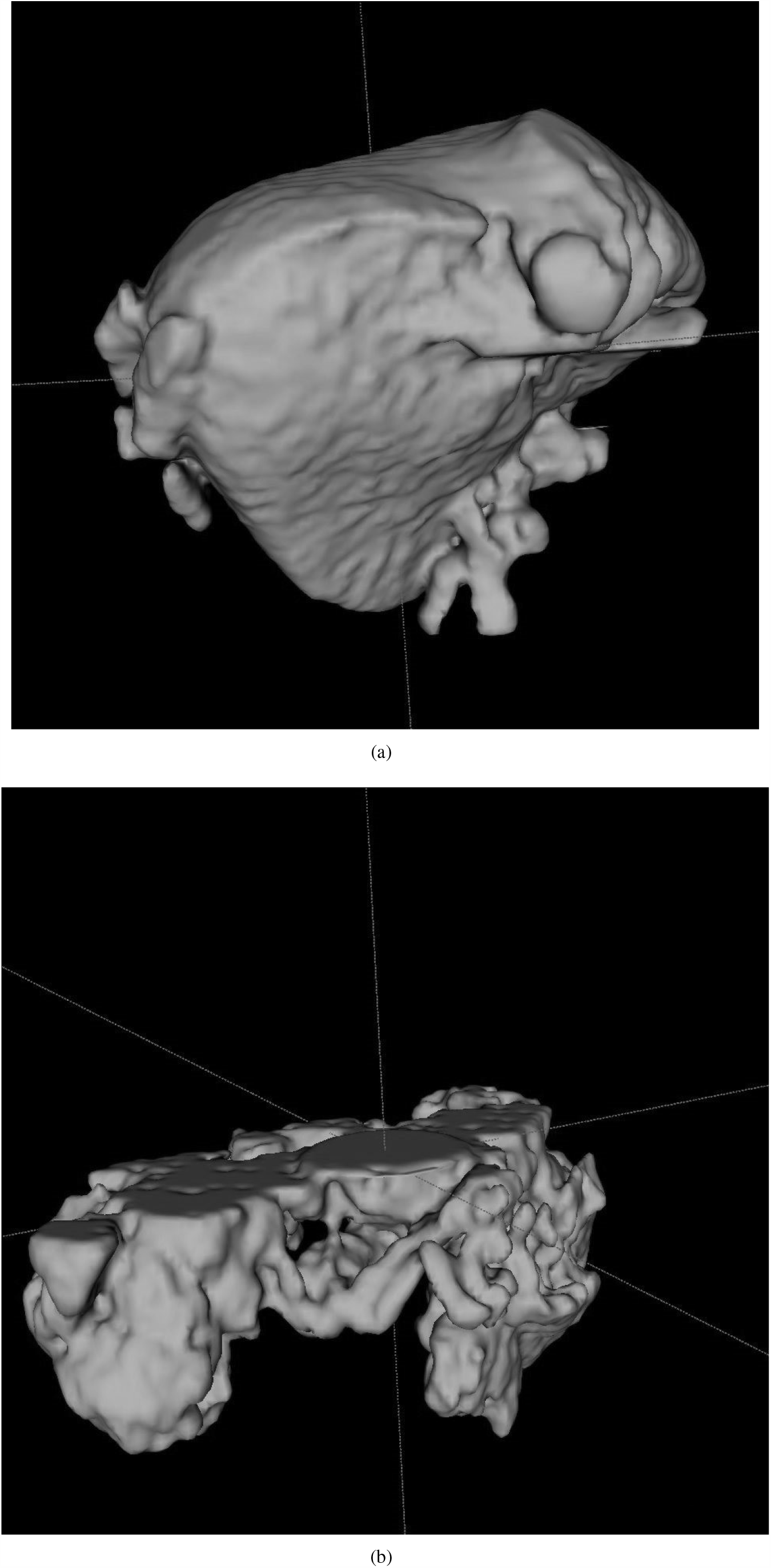
Example segmentations: (a) A good segmentation of the liver, with some leakage into surrounding structures. (b) A poor segmentation of the pancreas. The bounding box was shifted along the *z*-axis. This cut off the upper half of the pancreas, resulting in an incomplete segmentation. The segmentation also included parts of surrounding structures

Segmentation of unseen patient image data scan takes between 30 seconds and one minute depending on the size of the data, i.e. on an Intel-i7 mobile computer with an affordable NVIDIA GTX 1050 GPU (2-4 GB), which was too small to train 3D U-Nets.

In summary, the results of the current state of the segmentation method are affected by displaced bounding boxes and could not compete with the Dice coefficients of related studies. Though, it is important to mention that most of the studies had more training data and are often focused on a single organ not addressing a group.

## 4 Conclusion and Future Work

We were able to detect organ bounding boxes automatically. Nonetheless, the overall positioning and extension of the predicted bounding boxes have to be improved to ensure a satisfying segmentation.

We used predicted bounding boxes for a completely automatic method. Future works should focus on the accuracy and robustness of the BB detection. The quality of the target segmentation depends heavily on the quality of the detected bounding boxes.

## Data Availability

Data from public data bases as declared on p. 1

## Acknowledgements

This work was carried out at the Institute for Medical Informatics at the Universität zu Lübeck. DFG-MA 6791/1-1, Nvidia GPU grant, EXPLOR program by Stiftung Kessler + Co. für Bildung und Kultur.

